# Cost-effectiveness analysis of influenza vaccination with a high-dose vaccine versus an adjuvanted quadrivalent vaccine in older adults in Spain

**DOI:** 10.64898/2026.03.23.26349057

**Authors:** Jose María Abellán, Esther Redondo, Ángel Gil de Miguel, Iván Sanz Muñóz, Ariadna Diaz-Aguiló, Paloma Palomo, Manel Farré, Daniel Callejo, Marco Pinel, Juan Luis López-Belmonte

## Abstract

**Objectives:** Influenza is a widespread acute respiratory illness posing a major public health challenge for both the National Health Service (NHS) and society, particularly among older adults. This study aimed to assess the cost-effectiveness of high-dose quadrivalent vaccine (HD-QIV) versus adjuvanted quadrivalent vaccine (aQIV) in older adults in Spain.

**Methods:** Public health and economic benefits were evaluated using a decision-tree model considering influenza cases, GP and ED visits, hospitalizations, and influenza-related mortality. Deterministic and probabilistic sensitivity analyses addressed epidemiological and economic uncertainties.

**Results:** From a societal perspective, HD-QIV prevented 54,039 influenza cases, 7,733 GP consultations, 1,585 ED visits, 27,398 episodes of hospitalization due to cardiorespiratory events over a single influenza season and 1,203 deaths compared to aQIV when vaccinating adults ≥65 years old in Spain, resulting in 14,316 LYs and 12,440 QALYs gained over a lifetime horizon. The reduction in health outcomes outweighed the increase in vaccination costs, translating to a reduction in total costs with HD-QIV compared to aQIV. Therefore, vaccinating older adults in Spain with HD-QIV instead of aQIV was a dominant strategy when evaluating hospitalizations due to respiratory and cardiovascular events. HD-QIV remained dominant from a NHS perspective. Sensitivity analyses confirmed the robustness of the model.

**Conclusions:** This analysis showed that vaccinating older adults in Spain with HD-QIV instead of aQIV would reduce influenza cases, GP and ED visits, hospitalizations, deaths, and associated costs, and thus it should be the strategy of choice in a situation of budgetary constraints from either a societal or an NHS perspective.

**HIGHLIGHTS:**

- A decision-tree cost-effectiveness model addressed the evidence gap of lacking head-to-head trials between HD-QIV and aQIV in older adults, using the highest available evidence and indirect comparison via SD-QIV.
- HD-QIV prevented more influenza cases, hospitalizations, and deaths than aQIV, resulting in greater life-years and QALYs gained, and substantial cost savings for both society and the NHS.
- These findings support HD-QIV as the preferred vaccination strategy for older adults in Spain, providing robust evidence for policymakers to optimize influenza prevention and healthcare resource allocation.

## 1. Introduction

Seasonal influenza is an acute respiratory viral infectious disease that rapidly spreads during the winter season, leading to epidemics ^1^. In Spain, an averaged incidence of 793 cases of influenza per 100,000 inhabitants among older adults (≥65 years old) has been reported ^2^ and it is associated with a high clinical and economic burden ^1,3^. Adults aged ≥65 years are especially susceptible to influenza severe infections and related complications, and represent a substantial proportion of the burden of disease ^3^. Between 2008/09 and 2017/18, older adults represented 56.7% of hospitalizations with a lab-confirmed diagnosis of influenza, averaging 88.2 cases per 100,000 inhabitants, reaching up to 289.3 cases per 100,000 in the last season, with a mean case fatality risk of 7.0% ^4^. The mean annual cost of influenza hospitalizations in Spain was 45.7 million €, of which 52.8% was generated by adults ≥65 years old ^4^. In this population, influenza was also associated with 144.4 excess deaths per 100,000 inhabitants compared to 27.7 deaths per 100,000 for all age groups ^4^. These higher figures for the older adults can be explained through immunosenescence and the higher prevalence of comorbidities in this population ^5^.

Vaccination is the most effective strategy to prevent influenza disease and its complications ^6^. Health agencies in most European countries, including Spain, recommend and support seasonal influenza vaccination programs for high-risk populations, such as adults aged ≥60 or 65 years^7^.

Nevertheless, influenza vaccine effectiveness remains suboptimal, especially in the older adults. The high-dose (HD) influenza vaccine and the adjuvanted influenza vaccine were developed for its use in this age group. The HD influenza vaccine, which has four times more antigen (60 μg of hemagglutinin) per strain than a standard dose (SD) vaccine, has shown to provide better immunogenicity, efficacy and effectiveness than SD vaccines in adults ≥65, while keeping an acceptable safety profile ^8,9^. Adjuvanted vaccines contain a standard 15 μg dose of hemagglutinin for each of the four influenza strains included in the vaccine plus the MF59^®^ adjuvant, an oil-in-water emulsion of squalene oil ^10^. Adjuvanted vaccines have shown to provide higher immunogenicity than SD vaccines in adults ≥65 ^11^.

Currently, HD quadrivalent influenza vaccines (HD-QIV) and adjuvanted quadrivalent influenza vaccines (aQIV) are the most common vaccination strategies being used in Spain in older adults. The objective of the current study was to analyze the cost-effectiveness and burden of disease associated with vaccinating older adults with HD-QIV instead of aQIV in Spain.

## 2. Materials and methods

### 2.1. Model structure

A pharmacoeconomic model was used to explore the cost-effectiveness of HD-QIV compared to aQIV vaccination strategies for the Spanish population aged 65 years or older. This economic model was based on a decision-tree approach consistent with WHO guidance and requirements on the evaluation of influenza vaccinations ^12^, and has been published previously ^13–16^.

The decision-tree model structure is presented in Figure 1. The model estimates the hypothetical number of influenza cases in people ≥65 years using the influenza attack rate and vaccine efficacy. Individuals ≥65 years can be vaccinated or not; vaccinated individuals may receive HD-QIV or aQIV. The likelihood of laboratory-confirmed influenza (LCI) depends on the effectiveness of the vaccine. As no head-to-head trial compares HD-QIV and aQIV, the model uses each vaccine’s relative efficacy (rVE) against a common comparator, SD-QIV, to reduce bias from absolute efficacy (VE) across seasons and settings. Hospitalizations were estimated independently from influenza cases based on vaccine effectiveness in preventing them. Influenza-related hospitalizations were modeled through four definitions: pneumonia and influenza (P&I), respiratory cause, cardiorespiratory cause, or all-cause, with ICD10 codes in Supplementary Table 1. GP visits, ED visits, and influenza-related mortality were estimated independently, not mutually exclusive, and conditional on infection.

**Figure 1.**
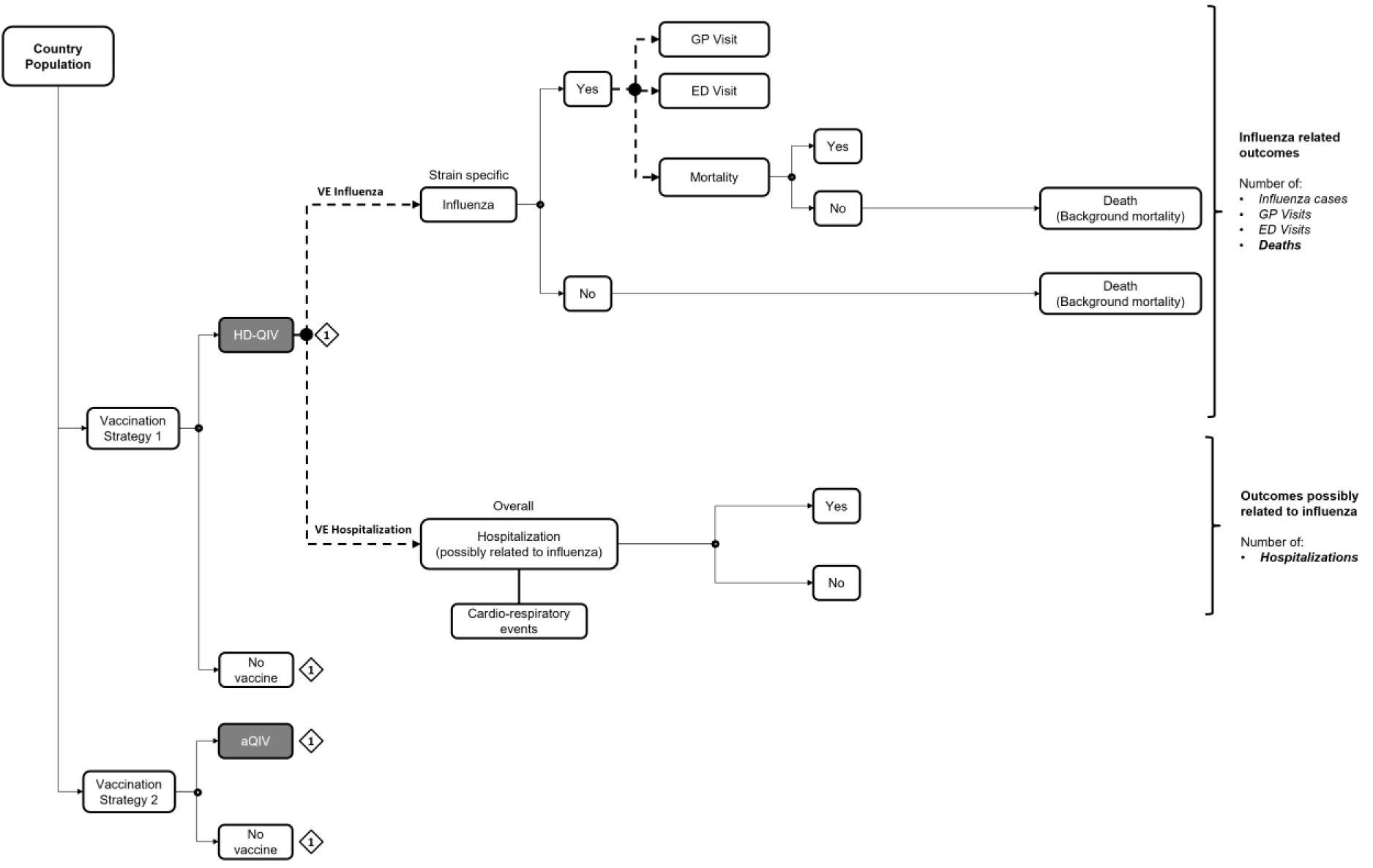
Model structure. The doted lines represent non mutually exclusive branches, while straight lines represent mutually exclusive branches. **Abbreviations:** aQIV, adjuvanted quadrivalent vaccine; ED, Emergency Department; GP, General Practitioner; HD-QIV, high dose quadrivalent vaccine; VE, vaccine effectiveness.

We assumed that all relevant events occurred within a single influenza season; therefore, a 6-month time horizon was considered, including episodes between Week 40 and Week 20 of the following year. Consequently, costs and health outcomes during this period did not require discounting. However, life-years (LYs) and quality-adjusted life years (QALYs) lost due to influenza-related premature deaths were discounted at 3.0%, in line with Spanish cost-effectiveness recommendations ^17^.

The base case analysis was conducted from a societal perspective, including direct medical costs (vaccine acquisition and administration), healthcare resource costs (GP and ED visits, hospitalizations), and indirect costs (productivity loss due to absenteeism and mortality). In a scenario analysis, indirect costs were excluded to adopt a healthcare perspective.

### 2.2. Model inputs

All inputs used throughout the model were obtained from the highest available level of evidence.

For the purpose of data granularity, two age groups were differentiated in the model: 65-74 years old, and ≥75 years old. Specific age-group data was used in the model whenever possible. If not available, the same value was applied for those aged 65 years and older.

The inputs used in the model are presented in Table 1.

**Table 1.**
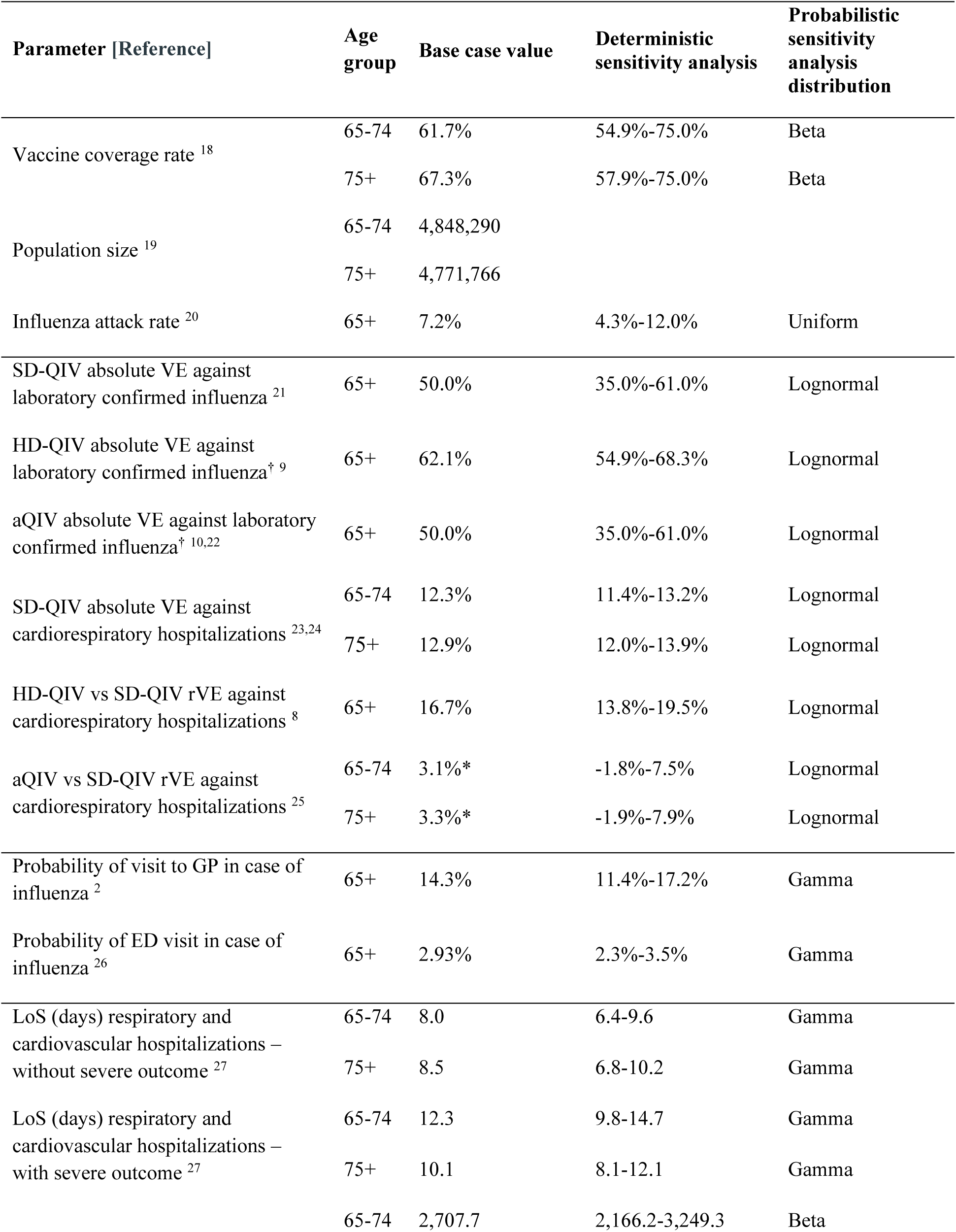
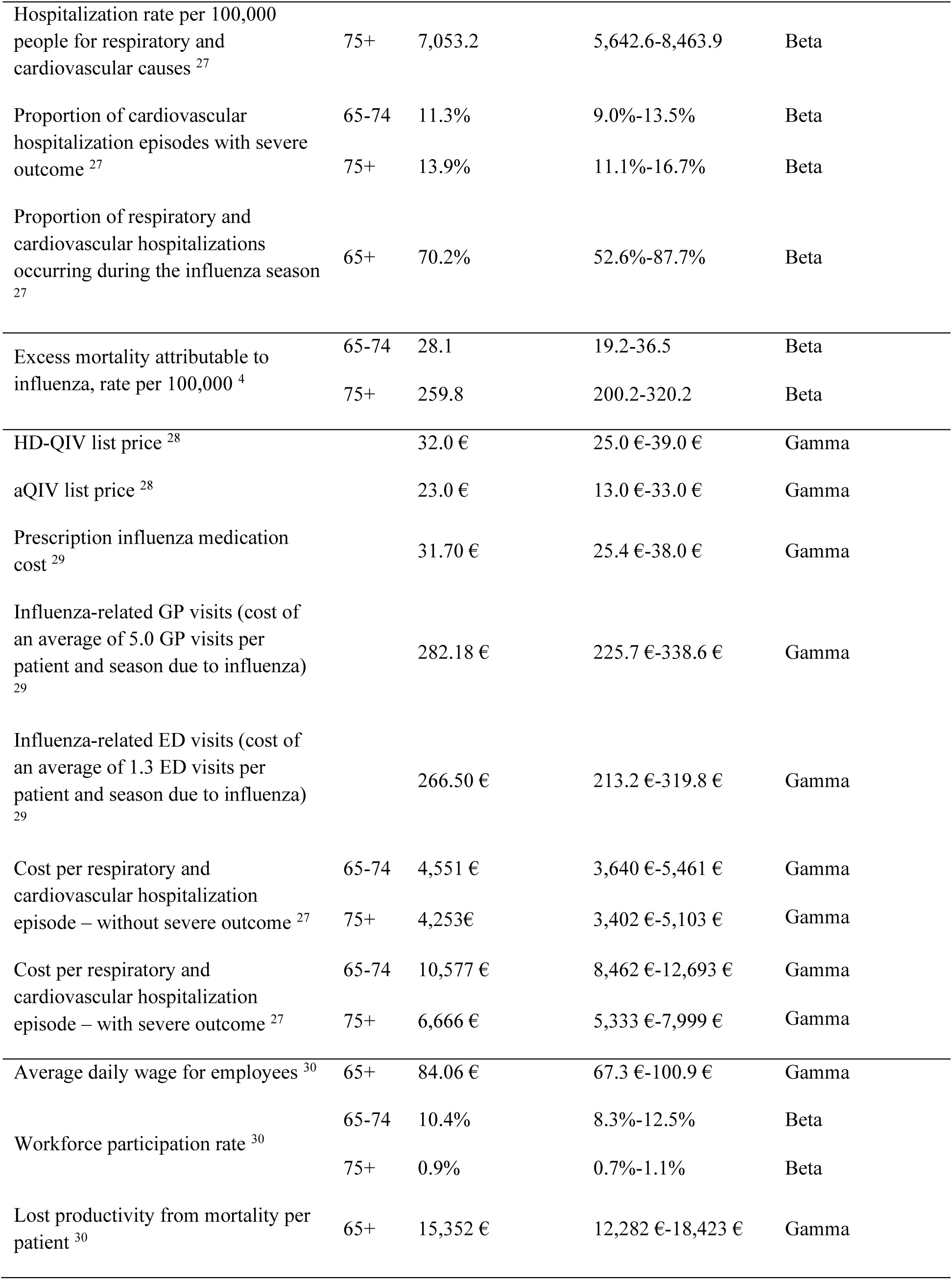
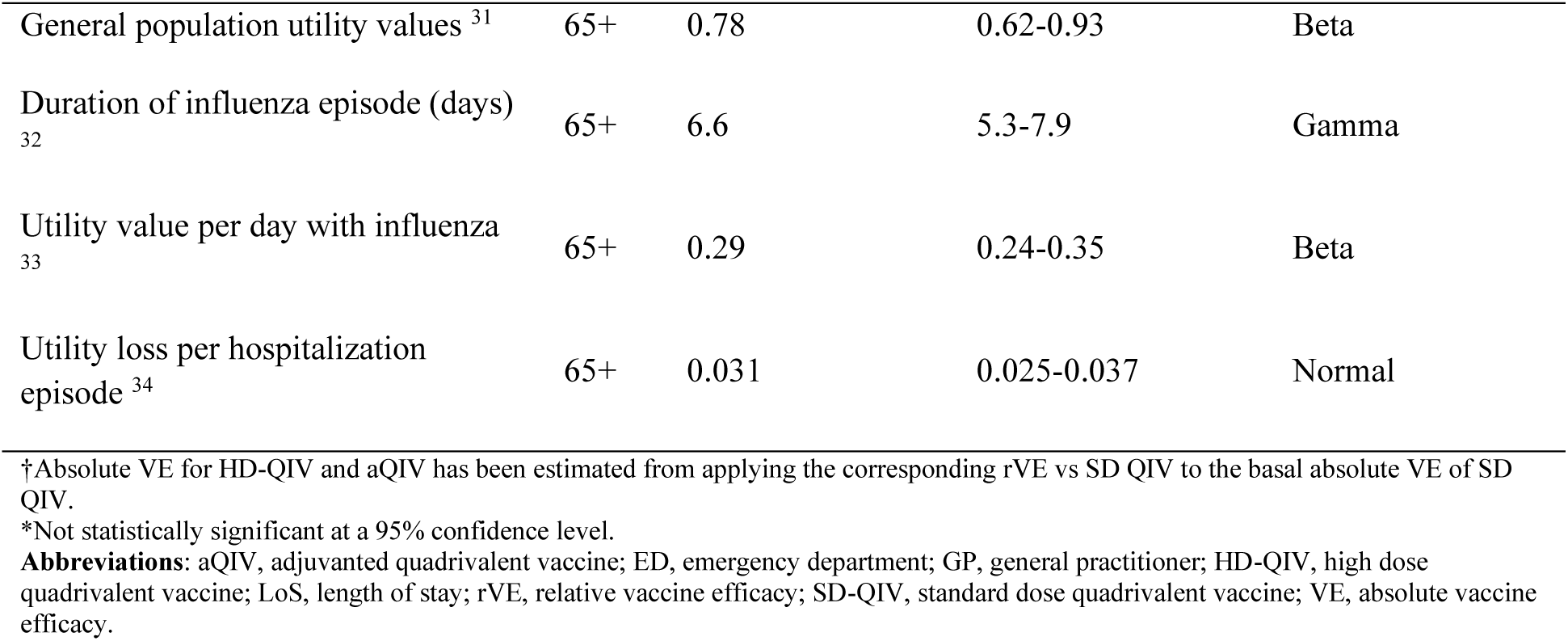
List of inputs included in the model.

#### 2.2.1. Population, attack rate & vaccine coverage

The number of target subjects to be vaccinated was obtained from the National Institute of Statistics of Spain for individuals aged ≥65 years ^19^. Influenza cases among this age group were informed by the attack rate from a systematic review and meta-analysis by Somes et al. (2018), representing the highest level of evidence. The pooled estimate of symptomatic attack rate from the controlled arm of three randomized trials (over 2,500 subjects) was 7.2% for older adults (≥65 years) ^20^. To account for vaccine uptake, the influenza coverage rate reported by the Spanish Ministry of Health (SIVAMIN) was applied ^18^, assuming equal coverage across vaccination strategies.

#### 2.2.2. Healthcare resource use

The probability of GP visits was estimated from the Surveillance System for Influenza and Other Respiratory Viruses in Spain ^2^, and the probability of at least one ED visit was derived from a regional influenza surveillance report ^26^. Both probabilities were conditional on influenza infection.

Hospitalization and mortality rates, as well as length of stay, were obtained from the Ministry of Health for the pre-COVID-19 period (2014–2019) ^27^. Hospitalizations leading to ICU admission or death were defined as severe outcomes, reflecting a worse health state ^27^. The percentage of hospitalizations with severe outcomes was sourced from the same period ^27^.

To reflect that not all cardiorespiratory hospitalizations occurred during the influenza season an adjustment factor was applied. Based on the Spanish Ministry of Health hospitalization database, 70.2% of these hospitalizations were assumed to occur within the influenza season ^27^.

#### 2.2.3. Vaccine efficacy against influenza cases

As previously mentioned, due to the absence of a head-to-head randomized trial between HD-QIV and aQIV, vaccine efficacy was estimated using a common comparator, SD-QIV. The VE of SD-QIV against laboratory-confirmed influenza was obtained from the randomized clinical trial (RCT) by Govaert et al. (1994), estimated at 50% ^21^.

Evidence on rVE for HD-QIV was inferred from the rVE of HD-TIV reported in the RCT by Diaz Granados et al. (2014) through an immunobridging exercise, resulting in an absolute efficacy of 62.1% for HD-QIV ^9,35,36^. For the adjuvanted vaccine, a systematic review by Murchu et al. (2022), including 48 RCTs and non-randomized studies, found no superiority over standard-dose vaccines, highlighting the lack of high-quality data ^22^. Additionally, a randomized clinical study showed that aQIV was not superior to no vaccination (VE: 19.8% [multiplicity-adjusted 95% CI −5.3 to 38.9] against all influenza) ^10^. Therefore, its absolute efficacy was assumed to be 50%, the same as SD-QIV.

#### 2.2.4. Vaccine efficacy against hospitalizations

VE for SD-QIV against respiratory hospitalizations, including pneumonia and influenza (P&I), was obtained from the Cochrane review by Beyer et al. (2013), where SD-TIV showed 28% efficacy against respiratory events; this was adjusted for SD-QIV to 29.3% ^23,24^. Both SD-TIV and SD-QIV demonstrated efficacy against cardiovascular (CV) events in high-risk CV patients ^37^, however, evidence for VE against CV hospitalizations in non-high-risk adults ≥65 years was lacking. Therefore, VE against respiratory events was weighted by the relative number of respiratory hospitalizations over total cardiorespiratory and all-cause hospitalizations from the Ministry of Health to estimate VE for these broader categories ^23^. SD-QIV was assumed to protect only against respiratory events (ICD10 codes J9-J18; J40-J45; J96).

The rVE for HD-QIV vs aQIV against hospitalizations was derived from the highest available evidence: a meta-analysis of RCTs (Skaarup et al., 2024) and a meta-analysis of RCTs and observational studies (Lee et al., 2023) ^8,38–40^. Skaarup et al. (2024) assessed rVE of HD-QIV vs SD-QIV in reducing P&I and all-cause hospitalizations in adults ≥65 years ^40^. Lee et al. (2023) analyzed over 12 influenza seasons and 45 million individuals ≥65 years, providing rVE estimates for HD-QIV vs SD-QIV against respiratory and cardiorespiratory hospitalizations ^8^.

For aQIV, the highest evidence identified was a cluster-randomized trial by McConeghy et al. (2021), which estimated rVE against P&I, respiratory, and all-cause hospitalizations ^25^. Cardiorespiratory hospitalizations were estimated using the same methodology as for SD-QIV, assuming aQIV only protected against respiratory events, despite this co-primary outcome not being statistically significant.

#### 2.2.5. Mortality

Mortality in the model was based on the excess mortality from influenza estimated for Spain. Therefore, the deaths prevented depend only on the influenza cases avoided with each vaccine and are not linked to hospitalizations. Mortality data was obtained from a real-world evidence study (the BARI study) that comprised ten epidemic seasons (from 2008/09 to 2017/18) and estimated the excess mortality in Spain on a national level ^4^.

#### 2.2.6. Utilities

To estimate QALYs, expected life years accrued by the model population were adjusted by age specific utility values for the Spanish general population ^31^. For subjects who develop influenza, a utility value specific to clinically defined influenza disease was applied daily for the assumed duration of the illness ^33^. An influenza episode was considered to last for 6.6 days, on average, as estimated from a systematic review and meta-analysis of randomized controlled trials ^32^. For subjects who experience an influenza-related hospitalization, QALY loss specific to this outcome was applied ^34^.

#### 2.2.7. Costs

List vaccine prices were considered in the model ^28^.

Costs per hospitalization episode, with or without severe outcome, for the defined hospitalization categories were obtained from an official Spanish database ^27^. Hospitalizations for each vaccination strategy were estimated, then the percentage with severe outcomes was applied to calculate episodes with or without severe outcome. These were multiplied by the corresponding unit cost.

Additionally, a retrospective cost-of-illness study in Spain informed costs of prescription influenza medications, GP visits, and ED presentations ^29^.

Indirect costs were included using workforce participation rates and average daily wages for employees aged ≥65 years, representing productivity loss from absenteeism due to influenza ^30^. Productivity loss from mortality was estimated using the friction cost method, imputing the average daily wage for six months, the assumed replacement period ^30^.

All costs are expressed in euros and were updated to 2023 through the Spanish Consumer Price Index published by the National Institute of Statistics ^41^.

### 2.3. Cost-effectiveness analysis

The cost-effectiveness analysis was conducted in Microsoft Excel^®^. Given that influenza can cause complications beyond respiratory infection, the cardiorespiratory definition was selected as the base case ^8,9,42^, as it better reflects the real impact of vaccination. The model estimated population-level health outcomes: influenza cases, GP and ED visits, hospitalizations, deaths during the influenza season, and LYs and QALYs over a lifetime horizon when HD-QIV or aQIV were used in individuals aged ≥65 years. Undiscounted societal costs and discounted QALYs were calculated for each strategy to derive the incremental cost-effectiveness ratio (ICER), defined as the difference in costs divided by the difference in health benefits between interventions.

As Spain lacks an official willingness-to-pay (WTP) threshold, €25,000/QALY was considered, corresponding to the upper limit estimated by Vallejo et al. (2018) ^43^ for the Spanish NHS (healthcare perspective) and Vallejo et al. (2020) for societal perspective ^44^. In line with recent updates, a WTP of 34,000 €/QALY has also been explored, following the updated threshold proposed by the same author ^45^.

Three alternative scenarios were also analyzed, considering influenza hospitalizations plus attributable pneumonia, respiratory outcomes, and all-cause hospitalizations (Supplementary Table 2).

Sensitivity analyses were performed to assess robustness. A univariate deterministic sensitivity analysis (DSA) used 95% confidence intervals (CI) for parameters when available or ±20% variation around base case values. Probabilistic sensitivity analysis (PSA) was conducted through 1,000 Monte Carlo simulations.

## 3. Results

In the base case, HD-QIV averted 54,039 influenza cases, 7,733 GP consultations, 1,585 ED visits, 27,398 episodes of hospitalization due to cardiorespiratory events and 1,203 deaths compared to aQIV when vaccinating adults ≥65 years old in Spain, which resulted in 14,316 LYs and 12,440 QALYs gained. The increased cost of vaccination with HD-QIV (Δ 55.8 million €) was offset by the savings on direct and indirect costs (Δ −154.0 million €), resulting in an absolute cost reduction of 98.2 million € (Table 2). Therefore, vaccinating older adults in Spain with HD-QIV instead of aQIV was a dominant strategy when evaluating hospitalizations due to respiratory and cardiovascular events.

**Table 2.**
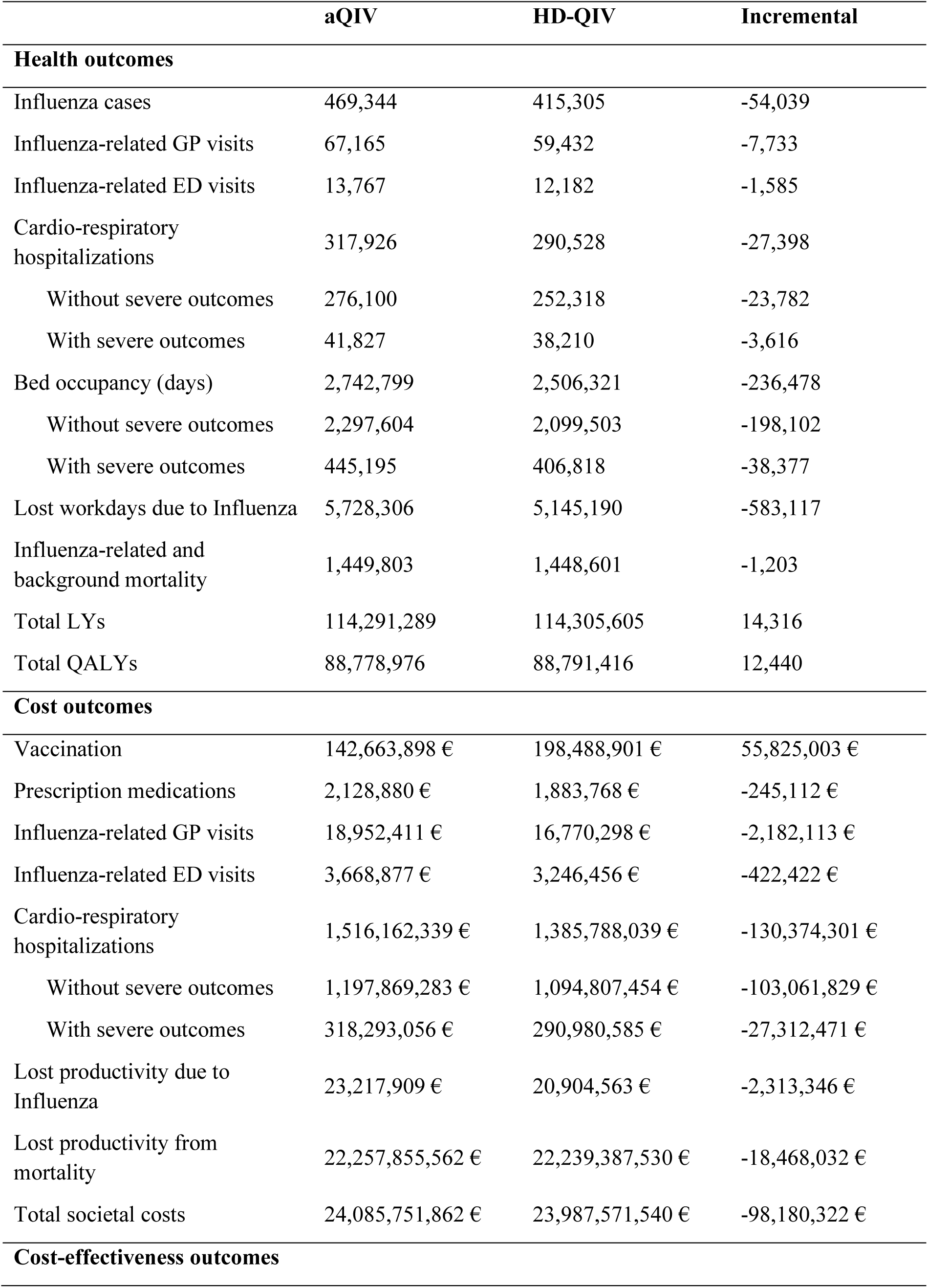
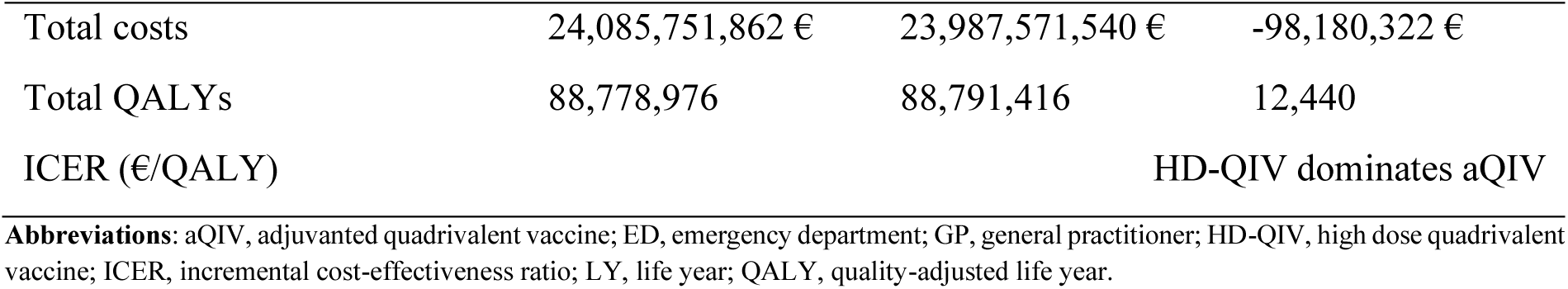
Clinical and economic model results for the base case: cardiorespiratory hospitalizations.

From the healthcare perspective, vaccinating with aQIV would lead to 1,804,678,391 € in total costs and 88,778,976 total QALYs; being 1,727,279,447 € and 88,791,416 for HD-QIV, respectively. This results in 77,398,944 € in savings and 12,440 incremental QALYs with HD-QIV, remaining the dominant strategy over aQIV.

The one-way DSA showed that the vaccines prices, the rVE of HD-QIV vs. SD-QIV against influenza cases, and the rVE of aQIV vs SD-QIV against hospitalizations were the parameters that most affected the results, although no variation in the parameters changed the conclusions of the analysis (Figure 2).

**Figure 2.**
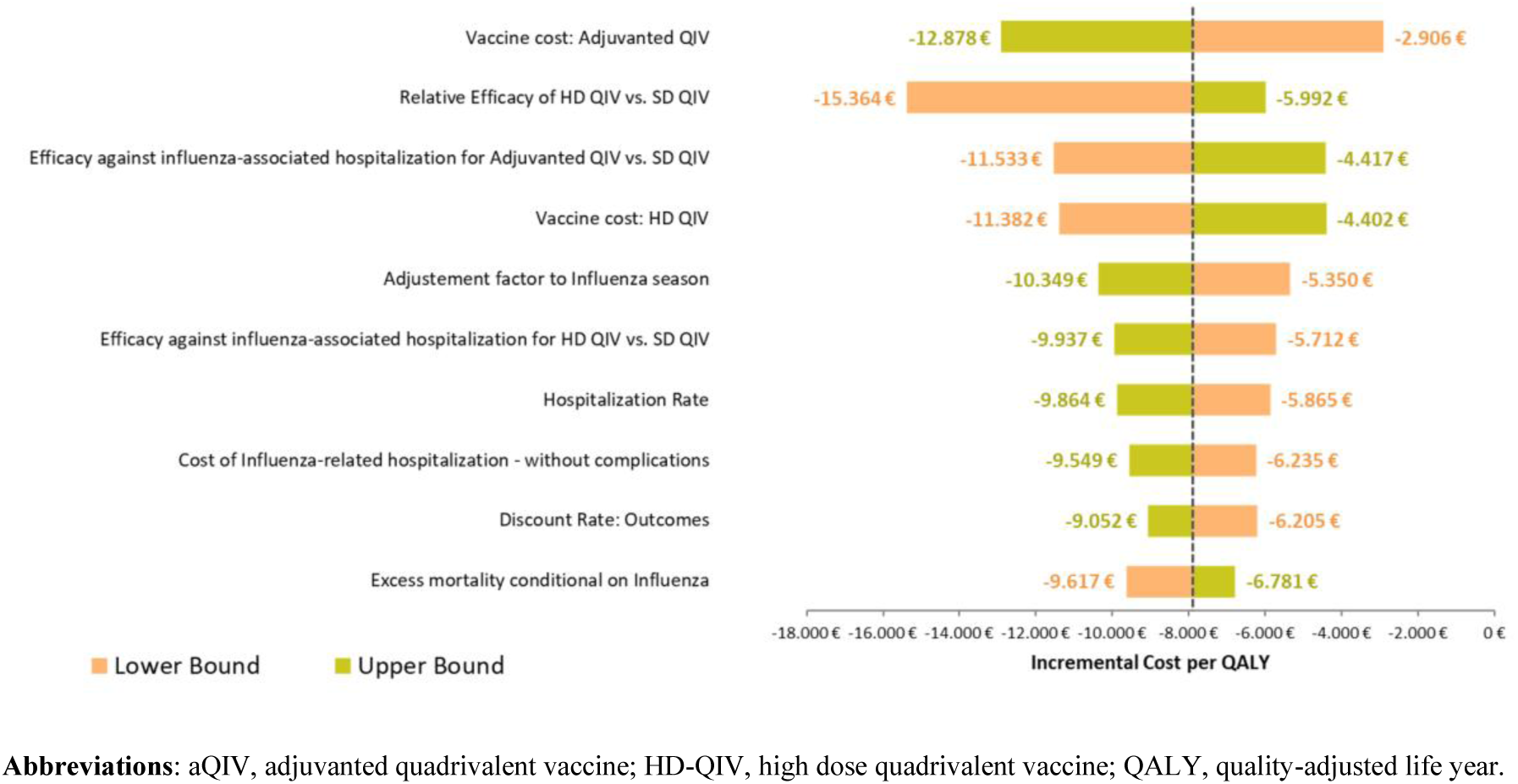
Deterministic sensitivity analysis for the base case: cardiorespiratory hospitalizations. **Abbreviations**: aQIV, adjuvanted quadrivalent vaccine; HD-QIV, high dose quadrivalent vaccine; QALY, quality-adjusted life year.

The PSA showed that for most of the iterations (989 out of the 1,000 [98.9%]), HD-QIV remained the dominant strategy compared to aQIV (Figure 3). For a WTP threshold of 25,000 €/QALY, the probability of HD-QIV being a cost-effective option for Spain was 100%, thus making the higher threshold of 34,000 €/QALY also cost-effective.

**Figure 3.**
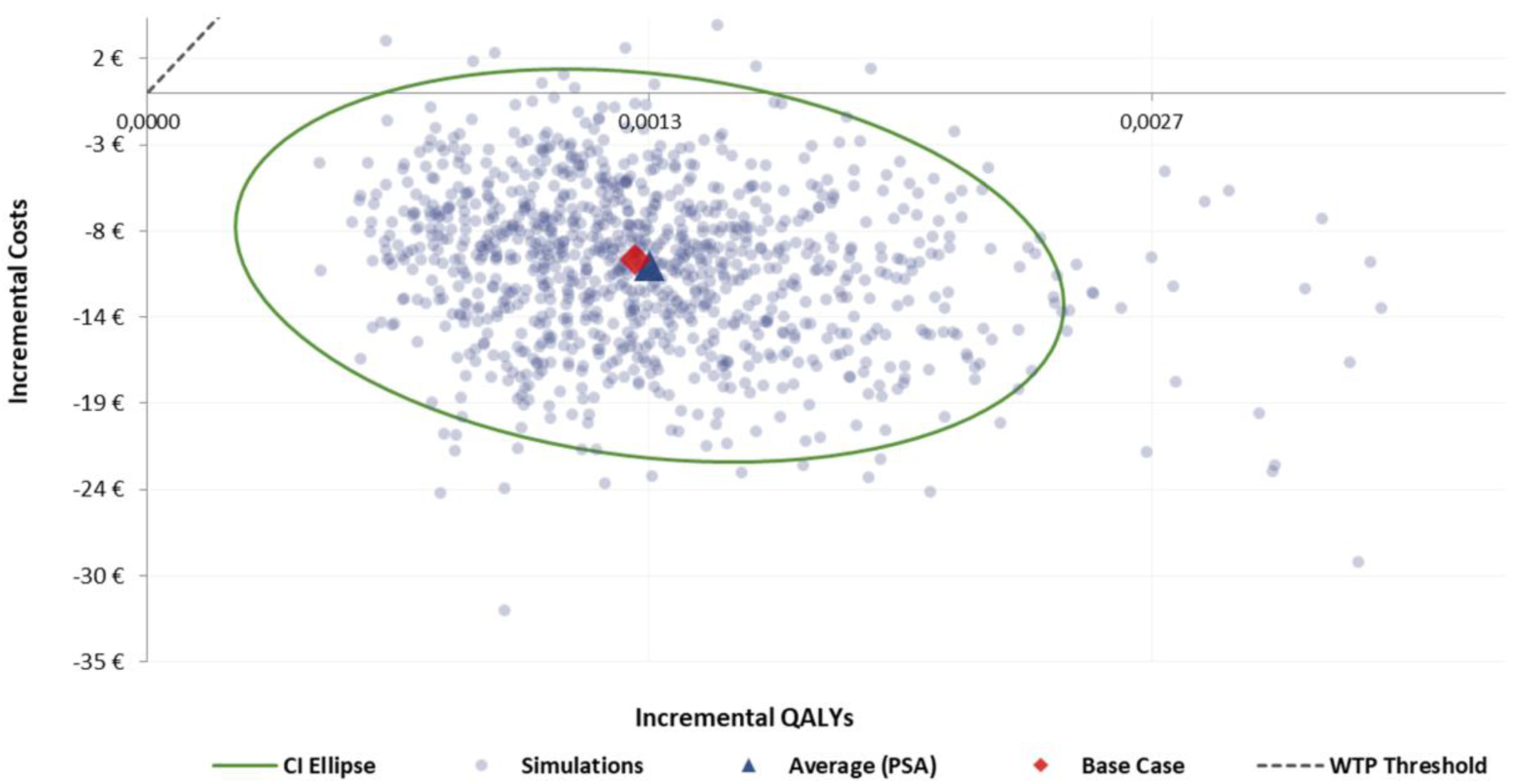
Cost-effectiveness plane for the base case: cardiorespiratory hospitalizations. **Abbreviations:** CI, confidence interval; PSA, probabilistic sensitivity analysis; QALY, quality-adjusted life year; WTP, willingness-to-pay.

In all the alternative scenarios, HD-QIV remained the cost-effective vaccination strategy compared to aQIV, showing dominancy in half of the scenarios evaluated (Supplementary Table 3).

## 4. Discussion

In our cost-effectiveness analysis from a societal perspective, vaccinating older adults (≥65 years) in Spain with HD-QIV was a dominant strategy compared to aQIV for preventing influenza cases, GP and ED visits, cardiorespiratory hospitalizations, and deaths, using the highest-quality evidence. For a single season, HD-QIV prevented 54,039 influenza cases, 7,733 GP visits, 1,585 ED visits, and 27,398 cardiorespiratory hospitalizations, freeing 236,478 bed-days. Reducing hospitalizations is particularly relevant for healthcare providers during high-demand winter seasons when resources are limited. The greater effectiveness of HD-QIV generated savings from fewer hospitalizations and healthcare visits, outweighing its higher vaccine cost. Additionally, reduced productivity loss contributed to total savings of €98.2 million over one season. From the healthcare perspective, HD-QIV saved €77.4 million. HD-QIV also prevented 1,203 deaths, translating into 12,440 QALYs and 14,316 LYs gained over a lifetime horizon compared to aQIV. Therefore, HD-QIV was not only more cost-effective but also more advantageous for the NHS from a budgetary perspective due to substantial savings.

The results of the sensitivity analysis showed that the dominance of HD-QIV was robust to changes in the model parameters. The key driver for the cost-effectiveness was the relative efficacy of HD-QIV vs SD-QIV, which responds to the model assumptions. Still, even when using the lower bound of the 95% CI for the rVE parameter value (as described in the Lee et al. meta-analysis ^8^), HD-QIV remained dominant vs aQIV. Other relevant parameters included rVE against hospitalizations for either HD-QIV and aQIV, and number of baseline hospitalizations.

In our study, a cardiorespiratory definition was used in the base case to assess vaccination effects on hospitalizations. Although less stringent than a P&I approach, growing evidence links influenza as a precipitating factor for destabilizing chronic conditions such as asthma or congestive heart failure, leading to complications beyond the respiratory domain ^8,9,42^. This approach captured severe outcomes potentially related to influenza that may not be identified in traditional surveillance networks. Moreover, the efficacy of HD-QIV vs SD-QIV against cardiorespiratory events has been evaluated in RCTs ^8,9^, supporting its selection as the base case. Scenario analysis confirmed HD-QIV remained cost-effective across all hospitalization definitions.

In accordance with reference-level-of-evidence hierarchies, systematic reviews and meta-analyses of randomized trials are considered the highest level of evidence, followed by individual RCTs, systematic reviews and meta-analyses not including randomized trials, cohort studies, case control studies, cross sectional surveys, and lastly case reports ^38,39^. For each kind of study, a systematic review is generally better than an individual study ^38,39^. This hierarchy has been used throughout our analysis to guarantee that the highest level of available evidence has been considered for each input of the model ^38,39^, even though the level of available evidence for HD-QIV and aQIV was not equal

The meta-analysis of RCTs and observational studies by Lee et al. (2023) was considered the highest level of evidence for the rVE of HD-QIV vs SD-QIV against P&I, respiratory, cardiorespiratory, and all-cause hospitalizations ^8^. For the adjuvanted vaccine, a non-targeted, non-systematic literature review was conducted to identify the highest-quality published evidence on aQIV effectiveness against hospitalizations; five publications were assessed ^22,25,46–48^. Coleman et al. (2021) reported a systematic review and meta-analysis of 21 non-interventional real-world studies comparing the adjuvanted vaccine with SD or HD vaccines, without RCTs, including posters/abstracts, and combining heterogeneous outcomes (hospitalizations and outpatient visits), yielding non-conclusive results ^46^. Domnich and de Waure (2022) synthesized 10 retrospective observational studies comparing aQIV with HD vaccines, concluding that evidence was inconclusive due to reliance on U.S. register-based data, restriction to non-LCI endpoints, limited seasons, small and variable pooled effects, and predominance of industry-sponsored studies ^48^. Murchu et al. (2022) included 48 RCTs and non-randomized studies and concluded the adjuvanted vaccine was not significantly different from non-adjuvanted vaccines, with small study numbers, statistical/clinical heterogeneity, and hospitalization outcomes not transparently reported (mixing absolute and relative measures); overall evidence was limited ^22^. Gartner et al. (2022) updated this review but combined influenza-related hospitalizations and ED visits, noted moderate-to-serious risk of bias, and very high heterogeneity (>95%), making the outcomes unsuitable for our model ^47^. McConeghy et al. (2021) conducted a cluster-randomized clinical trial in U.S. nursing homes during the 2016/17 season, randomizing >50,000 residents to adjuvanted vs non-adjuvanted vaccines, with outcomes including P&I, respiratory, and all-cause hospitalizations ^25^. P&I and all-cause hospitalizations were statistically significant with hazard ratio confidence intervals close to 1, while respiratory hospitalizations were not significant (p = 0.19) ^25^. Given that the systematic reviews provided lower-quality or non-applicable evidence for hospitalization outcomes, McConeghy et al. (2021) was considered the best available evidence to inform aQIV effectiveness against hospitalizations. Nonetheless, this trial had limitations: i) a restricted sample that may not represent older adults in the general population; ii) nursing home residents in the United States may not reflect residents in Spain; and iii) data were limited to a single influenza season ^25^.

Our study is not the first one to assess the cost-effectiveness of HD-QIV in older adults in Spain. Redondo et al. (2021) evaluated the cost-effectiveness of HD-QIV vs an adjuvanted trivalent vaccine and determined that HD-QIV is a cost-effective strategy in Spain to vaccinate older adults reducing cases of influenza, GP visits, hospitalizations (from either a cardiorespiratory or an influenza broad definition), deaths, and associated healthcare costs ^15^. In turn, Ruiz-Aragón et al. (2022) compared HD-QIV to aQIV and concluded that aQIV provided a highly cost-effective alternative to HD-QIV for the vaccination of people aged 65 years or older in Spain ^49^. However, this analysis holds limitations that impact results and should be considered for interpretation. First, assumptions made on the rVE of aQIV and HD-QIV may be affected by biases that are not clearly resolved. Economic evaluations of vaccines are highly sensitive to the efficacy assumptions used. Authors derived rVE estimates from a meta-analysis of eight studies that, to date, is neither published nor peer-reviewed, and its methods are unknown, with a high heterogeneity in rVE estimates (I^2^ = 92%) ^49^. Out of the eight studies, four were identified by a non-systematic targeted review without a proper risk of bias assessment, which is an important limitation given that all the studies included were observational and therefore subject to biases ^49^; while the other four studies came from the systematic review and meta-analysis performed by Coleman et al., susceptible to other methodological issues previously mentioned ^46,49^. Second, the authors assumed a comparable performance between HD-QIV and aQIV without mentioning that aQIV did not show efficacy vs a non-influenza vaccine in a RCT ^10^. Last, authors’ comparison between HD-QIV and aQIV differ from independent evaluations carried out in Europe and Spain using well-recognized quality appraisal frameworks ^22^. These evaluations highlighted the lack of robust data comparing adjuvanted vs. non-adjuvanted vaccines, while confirming the improved protection provided by high-dose vaccines vs standard dose and the robustness of evidence ^22^.

We believe our study overcomes these limitations by informing model inputs with the highest level of available evidence, providing a clear and detailed methodological description, and transparently justifying data sources used which are publicly available.

Nevertheless, our analysis is also subject to some limitations. The first limitation is that given the lack of randomized head-to-head studies, the effectiveness between HD-QIV and aQIV had to be indirectly compared through a common comparator, SD-QIV, an approach that has been used previously ^14–16^. Another limitation is that due to the lack of evidence comparing HD-QIV to SD-QIV, it was assumed that the prevention of influenza cases was identical to that of HD-TIV vs. SD-TIV. This assumption was deemed reasonable since the inclusion of a second B strain in the quadrivalent vaccine does not imperil protection against other strains, and therefore rVE should remain unaltered, as suggested by an immunobridging study and approved by the European Medicines Agency (EMA) ^35,36^. The third limitation is that, due to the limited body of evidence for aQIV, it was assumed that aQIV provided no additional benefit to SD-QIV in preventing influenza cases and cardiovascular hospitalizations ^22^. Even though aQIV has shown to have a higher immunogenicity than SD-QIV, it should be noted that although multiple studies on influenza infection suggest that hemagglutination inhibition (HI) antibody titers of 1:40 or higher correlate with immunity to influenza illness, HI titers do not serve as an absolute surrogate marker in the sense there is no universally agreed-upon threshold titer that defines clinical protection, as has been established by the EMA ^50^. Furthermore, we used a static model, instead of a dynamic transmission model, which for an infectious disease could have underestimated the value of the vaccine. Although this approach does not consider potential indirect benefits of vaccinations, the WHO guidelines suggest that decision trees are a suitable approach for evaluating costs and outcomes of an intervention within a limited timeframe, and also recommended for targeting the older adults ^12^. Lastly, inputs used in the model were taken from different public sources and published studies, with different designs. This limitation is inherent to most pharmacoeconomic models and was minimized through being consistent with data sources (e.g. public data was retrieved from the Ministry of Health whenever possible, data from BARI study was prioritized) and time periods.

## 5. Conclusions

In the context of implementing influenza immunization programs for older adults, it is important to consider other factors beyond the cost of the vaccine, such as promoting healthy aging, minimizing resource utilization and costs related to influenza cases, preventing respiratory and cardiovascular hospitalizations, and avoiding overload on healthcare services. Consequently, in Spain HD-QIV is a dominant strategy in influenza prevention for the older adults, compared to aQIV, by preventing influenza cases, GP visits, cardiorespiratory hospitalizations, deaths, and associated healthcare costs.

## 6. Declarations

### Author contribution

All authors contributed to the study conception and design. Material preparation, data collection and analysis were performed by Ariadna Diaz-Aguiló, Paloma Palomo, Manel Farré, Daniel Callejo, Marco Pinel, and Juan Luis López-Belmonte. The first draft of the manuscript was written by Daniel Callejo and Marco Pinel and all authors commented on previous versions of the manuscript. All authors read and approved the final manuscript.

### Data availability

No new datasets were generated or analyzed for this study. All information necessary to reproduce the analyses, including model structure, assumptions, and literature sources for all inputs, is reported in the article. The model code/executable files are proprietary and are not publicly available; aggregated, non-proprietary outputs can be provided by the corresponding author upon reasonable request.

### Ethical approval and informed consent

This study does not involve real patients; instead, it is based on a simulated cohort of patients. Therefore, ethical approval and informed consent are not applicable.

## References

1. Pérez-Rubio A, Platero L, Eiros Bouza JM. Seasonal influenza in Spain: Clinical and economic burden and vaccination programmes. Medicina Clínica (English Edition) 2019; 153(1): 16–27.

2. Informes anuales entre las temporadas 2011/12 y la 2019/20. Sistema de Vigilancia de la Gripe y otros virus respiratorios en España. Instituto de Salud Carlos III..

3. Oliva J, Delgado-Sanz C, Larrauri A, System tSIS. Estimating the burden of seasonal influenza in Spain from surveillance of mild and severe influenza disease, 2010-2016. Influenza and Other Respiratory Viruses 2018; 12(1): 161–70.

4. Pumarola T, Díez-Domingo J, Martinón-Torres F, et al. Excess hospitalizations and mortality associated with seasonal influenza in Spain, 2008-2018. BMC Infect Dis 2023; 23(1): 86.

5. Crooke SN, Ovsyannikova IG, Poland GA, Kennedy RB. Immunosenescence: A systems-level overview of immune cell biology and strategies for improving vaccine responses. Experimental Gerontology 2019; 124: 110632.

6. WHO. Fact sheet: seasonal influenza [Internet]. 2014 http://www.who.int/mediacentre/factsheets/fs211/en/.

7. Ministerio de Sanidad Consumo y Bienestar Social. Recomendaciones Vacunación Frente a la Gripe: Temporada 2023–2024. Available online: https://www.sanidad.gob.es/areas/promocionPrevencion/vacunaciones/programasDeVacunacion/docs/Recomendaciones_vacunacion_gripe.pdf (accessed on September 2023).

8. Lee JKH, Lam GKL, Yin JK, Loiacono MM, Samson SI. High-dose influenza vaccine in older adults by age and seasonal characteristics: Systematic review and meta-analysis update. Vaccine: X 2023; 14: 100327.

9. DiazGranados CA, Dunning AJ, Kimmel M, et al. Efficacy of High-Dose versus Standard-Dose Influenza Vaccine in Older Adults. New England Journal of Medicine 2014; 371(7): 635–45.

10. Beran J, Reynales H, Poder A, et al. Prevention of influenza during mismatched seasons in older adults with an MF59-adjuvanted quadrivalent influenza vaccine: a randomised, controlled, multicentre, phase 3 efficacy study. Lancet Infect Dis 2021; 21(7): 1027–37.

11. Verma SK, Mahajan P, Singh NK, et al. New-age vaccine adjuvants, their development, and future perspective. Frontiers in Immunology 2023; 14.

12. Newall AT, Chaiyakunapruk N, Lambach P, Hutubessy RCW. WHO guide on the economic evaluation of influenza vaccination. Influenza Other Respir Viruses 2018; 12(2): 211–9.

13. Alvarez FP, Chevalier P, Borms M, et al. Cost-effectiveness of influenza vaccination with a high dose quadrivalent vaccine of the elderly population in Belgium, Finland, and Portugal. Journal of Medical Economics 2023; 26(1): 710–9.

14. Mattock R, Gibbons I, Moss J, et al. Cost-effectiveness of high dose versus adjuvanted trivalent influenza vaccines in England and Wales. Journal of Medical Economics 2021; 24(1): 1261–71.

15. Redondo E, Drago G, López-Belmonte JL, et al. Cost-utility analysis of influenza vaccination in a population aged 65 years or older in Spain with a high-dose vaccine versus an adjuvanted vaccine. Vaccine 2021; 39(36): 5138–45.

16. Rumi F, Basile M, Cicchetti A, Alvarez FP, Azzi MV, Muzii B. Cost-effectiveness for high dose quadrivalent versus the adjuvanted quadrivalent influenza vaccine in the Italian older adult population. Front Public Health 2023; 11: 1200116.

17. López Bastida J, Oliva J, Antoñanzas F, et al. Propuesta de guía para la evaluación económica aplicada a las tecnologías sanitarias. Gaceta Sanitaria 2010; 24(2): 154–70.

18. SIVAMIN. Cobertura de vacunación por gripe 2022. Ministerio de Sanidad. Gobierno de España.

19. Instituto Nacional de Estadística. Población residente por fecha, sexo y edad. Datos provisionales a 01/07/2022. Accedido 22/05/2023.

20. Somes MP, Turner RM, Dwyer LJ, Newall AT. Estimating the annual attack rate of seasonal influenza among unvaccinated individuals: A systematic review and meta-analysis. Vaccine 2018; 36(23): 3199–207.

21. Govaert TM, Thijs CT, Masurel N, Sprenger MJ, Dinant GJ, Knottnerus JA. The efficacy of influenza vaccination in elderly individuals. A randomized double-blind placebo-controlled trial. Jama 1994; 272(21): 1661–5.

22. E OM, Comber L, Jordan K, et al. Systematic review of the efficacy, effectiveness and safety of MF59(®) adjuvanted seasonal influenza vaccines for the prevention of laboratory-confirmed influenza in individuals ≥18 years of age. Rev Med Virol 2023; 33(3): e2329.

23. Beyer WEP, McElhaney J, Smith DJ, Monto AS, Nguyen-Van-Tam JS, Osterhaus ADME. Cochrane re-arranged: Support for policies to vaccinate elderly people against influenza. Vaccine 2013; 31(50): 6030–3.

24. Chit A, Roiz J, Briquet B, Greenberg DP. Expected cost effectiveness of high-dose trivalent influenza vaccine in US seniors. Vaccine 2015; 33(5): 734–41.

25. McConeghy KW, Davidson HE, Canaday DH, et al. Cluster-randomized Trial of Adjuvanted Versus Nonadjuvanted Trivalent Influenza Vaccine in 823 US Nursing Homes. Clin Infect Dis 2021; 73(11): e4237–e43.

26. Dirección General de Salud Pública, Gobierno de Aragón. Informe de vigilancia de la temporada gripal. Aragón. 2017-2018. http://wwwaragones/DepartamentosOrganismosPublicos/Departamentos/Sanidad/AreasTematicas/SanidadProfesionales/SaludPublica/VigilanciaEpidemiologica/RedCentinela/ci03_Vigilancia_de_la_gripedetalleDepartamento.

27. Ministerio de Sanidad. Explotación de bases del CMBD. Estadísticos de referencia estatal de los sistemas de agrupación de registro de pacientes. Gobierno de España.

28. Vademecum [20 october 2023]. Available online: https://www.vademecum.es/

29. Gil-de-Miguel Á, Martinón-Torres F, Díez-Domingo J, et al. Clinical and economic burden of physician-diagnosed influenza in adults during the 2017/2018 epidemic season in Spain. BMC Public Health 2022; 22(1): 2369.

30. Instituto Nacional de Estadística, Encuesta anual de estructura salarial. 2020.

31. Crépey P, Redondo E, Díez-Domingo J, Ortiz de Lejarazu R, Martinón-Torres F, Gil de Miguel Á, et al. (2020) From trivalent to quadrivalent influenza vaccines: Public health and economic burden for different immunization strategies in Spain. PLoS ONE 15(5): e0233526. 10.1371/journal.pone.0233526.

32. Kuo Y-C, Lai C-C, Wang Y-H, Chen C-H, Wang C-Y. Clinical efficacy and safety of baloxavir marboxil in the treatment of influenza: A systematic review and meta-analysis of randomized controlled trials. *Journal of Microbiology*, Immunology and Infection 2021; 54(5): 865–75.

33. Turner D, Wailoo A, Nicholson K, Cooper N, Sutton A, Abrams K. Systematic review and economic decision modelling for the prevention and treatment of influenza A and B. Health Technol Assess 2003; 7(35): iii-iv, xi-xiii, 1–170.

34. Hollmann M, Garin O, Galante M, Ferrer M, Dominguez A, Alonso J. Impact of Influenza on Health-Related Quality of Life among Confirmed (H1N1)2009 Patients. PLOS ONE 2013; 8(3): e60477.

35. College Ter Beoordeling Van Geneesmiddelen (CBG MEB). Public Assessment Report. Scientific disucussion. Efluelda, suspension for injection in pre-filled syringe. NL/H/4757/001/DC. Available at: https://mri.cts-mrp.eu/Human/Product/Details/59731 (last access January 2021). 2020.

36. Chang L-J, Meng Y, Janosczyk H, Landolfi V, Talbot HK, Group QS. Safety and immunogenicity of high-dose quadrivalent influenza vaccine in adults 65 years of age: A phase 3 randomized clinical trial. Vaccine. 2019;37:5825–34.

37. Fröbert O, Götberg M, Erlinge D, et al. Influenza Vaccination After Myocardial Infarction: A Randomized, Double-Blind, Placebo-Controlled, Multicenter Trial. Circulation 2021; 144(18): 1476–84.

38. Burns PB, Rohrich RJ, Chung KC. The levels of evidence and their role in evidence-based medicine. Plast Reconstr Surg 2011; 128(1): 305–10.

39. OCEBM Levels of Evidence Working Group*. “The Oxford Levels of Evidence 2”. Oxford Centre for Evidence-Based Medicine. https://www.cebm.ox.ac.uk/resources/levels-of-evidence/ocebm-levels-of-evidence.

40. Skaarup KG, Lassen MCH, Modin D, et al. The relative vaccine effectiveness of high-dose vs standard-dose influenza vaccines in preventing hospitalization and mortality: A meta-analysis of evidence from randomized trials. Journal of Infection 2024; 89(1): 106187.

41. Instituto Nacional de Estadística. Indice de Precios de Consumo. Base 2021.

42. Net P, Colrat F, Nascimento Costa M, Bianic F, Thommes E, Alvarez FP. Estimating public health and economic benefits along 10 years of Fluzone® High Dose in the United States. Vaccine 2021; 39: A56–A69.

43. Vallejo-Torres L, García-Lorenzo B, Serrano-Aguilar P. Estimating a cost-effectiveness threshold for the Spanish NHS. Health Economics 2018; 27(4): 746–61.

44. Vallejo-Torres L, García-Lorenzo B, Rivero-Arias O, Pinto-Prades JL. The societal monetary value of a QALY associated with EQ-5D-3L health gains. The European Journal of Health Economics 2020; 21(3): 363–79.

45. Vallejo-Torres L. Estimating the Incremental Cost Per QALY Produced by the Spanish NHS: A Fixed-Effect Econometric Approach. Pharmacoeconomics 2025; 43(1): 109–22.

46. Coleman BL, Sanderson R, Haag MDM, McGovern I. Effectiveness of the MF59-adjuvanted trivalent or quadrivalent seasonal influenza vaccine among adults 65 years of age or older, a systematic review and meta-analysis. Influenza Other Respir Viruses 2021; 15(6): 813–23.

47. Gärtner BC, Weinke T, Wahle K, et al. Importance and value of adjuvanted influenza vaccine in the care of older adults from a European perspective – A systematic review of recently published literature on real-world data. Vaccine 2022; 40(22): 2999–3008.

48. Domnich A, de Waure C. Comparative effectiveness of adjuvanted versus high-dose seasonal influenza vaccines for older adults: a systematic review and meta-analysis. International Journal of Infectious Diseases 2022; 122: 855–63.

49. Ruiz-Aragón J, Márquez-Peláez S, Gani R, Alvarez P, Guerrero-Luduena R. Cost-Effectiveness and Burden of Disease for Adjuvanted Quadrivalent Influenza Vaccines Compared to High-Dose Quadrivalent Influenza Vaccines in Elderly Patients in Spain. Vaccines 2022; 10(2): 176.

50. European Medicines Agency (EMA). Fluad Tetra. Assessment report Procedure No. EMEA/H/C/004993/0000.

51. DiazGranados CA, Robertson CA, Talbot HK, Landolfi V, Dunning AJ, Greenberg DP. Prevention of serious events in adults 65 years of age or older: A comparison between high-dose and standard-dose inactivated influenza vaccines. Vaccine 2015; 33(38): 4988–93

