## Supplementary Table for "Cost-effectiveness analysis of influenza vaccination with a high-dose vaccine versus an adjuvanted quadrivalent vaccine in older adults in Spain"

### Supplementary Material

**Supplementary Table 1.** ICD10 codes considered in each of the definitions of hospitalization contemplated in the model

| **Hospitalization definition** | | **ICD10 codes** |
| --- | --- | --- |
| Cardiorespiratory hospitalization | | J09-J18, J40-45, J96, I20, I21, I24, I25, I50, I60-I63; I65, I66, I69, G45, G46 |
| Influenza & pneumonia | | J09, J10, J11, J12, J13, J14, J15, J16, J17,J18 |
| Respiratory hospitalization | | J09-J18, J40-45, J96 |
| All-cause hospitalization | | All codes |
| **ICD10 code** | **Category** | **Definition** |
| J09 | Pneumonia & influenza | Influenza due to identified influenza viruses |
| J10 |  | Flu due to other types of influenza viruses identified |
| J11 |  | Flu due to unidentified influenza viruses |
| J12 |  | Viral pneumonia, not classified elsewhere |
| J13 |  | *Streptococcus pneumoniae* pneumonia |
| J14 |  | *Haemophilus influenzae* pneumonia |
| J15 |  | Bacterial pneumonia, not otherwise classifiable |
| J16 |  | Pneumonia due to other infectious microorganisms, not classified elsewhere |
| J17 |  | Pneumonia in diseases classified elsewhere |
| J18 |  | Pneumonia, unspecified organism |
| J40 | Respiratory disease | Bronchitis, acute or chronic (not specified) |
| J41 |  | Simple and mucopurulent chronic bronchitis |
| J42 |  | Unspecified chronic bronchitis |
| J43 |  | Emphysema |
| J44 |  | Other chronic obstructive lung diseases |
| J45 |  | Asthma |
| J96 |  | Respiratory failure, not classified elsewhere |
| G45 | Cardiovascular disease | Transient ischemic strokes and related syndromes |
| G46 |  | Cerebral vascular syndromes in cerebrovascular diseases |
| I20 |  | Angina pectoris |
| I21 |  | Acute myocardial infarction with ST elevation (STEMI) and non-ST elevation (STEMI) |
| I24 |  | Other acute ischemic heart diseases |
| I25 |  | Chronic ischemic heart disease |
| I50 |  | Heart failure |
| I60 |  | Nontraumatic subarachnoid hemorrhage |
| I61 |  | Nontraumatic intracerebral hemorrhage |
| I62 |  | Other nontraumatic intracranial hemorrhage and those unspecified |
| I63 |  | Cerebral stroke |
| I65 |  | Occlusion and stenosis of precerebral arteries, which does not produce cerebral infarction |
| I66 |  | Occlusion and stenosis of cerebral arteries, which does not produce cerebral infarction |
| I69 |  | Cerebrovascular disease sequelae |

**Note**: The ICD10 codes for each definition are based on and converted from the MedDRA preferred terms for serious adverse events taken from a supplementary analysis of the randomized clinical trial (RCT) FIM12 ^51^.

**Supplementary Table 2.** Scenarios analyzed in the economic evaluation

| **Parameter** | **Age group** | **P&I hosp.** | **Respiratory hosp.** | **All cause hosp.** |
| --- | --- | --- | --- | --- |
| SD-QIV VE against hospitalizations ^23,24^ | 65-74 | 29.3% | 29.3% | 2.0% |
|  | 75+ | 29.3% | 29.3% | 3.1% |
| HD-QIV vs SD-QIV rVE against hospitalizations ^8^ | 65+ | 23.5% | 14.7% | 7.3% |
| aQIV vs SD-QIV rVE against hospitalizations | 65+ | 0% - 20.0% | 0% - 7.0% | 0% - 6.0% |
| Proportion of hosp. occurring during the influenza season ^27^ | 65+ | 100% | 74% | 66% |
| Hospitalization rate per 100,000 people ^27^ | 65-74 | 457.9 | 1,133.58 | 16,305.58 |
|  | 75+ | 1,560.0 | 3,108.35 | 29,131.60 |
| LoS (days) hospitalizations – without severe outcome ^27^ | 65-74 | 7.7 | 7.9 | 7.1 |
|  | 75+ | 8.5 | 8.3 | 8.4 |
| LoS (days) hospitalizations – with severe outcome ^27^ | 65-74 | 16.0 | 15.4 | 14.9 |
|  | 75+ | 10.6 | 10.6 | 11.9 |
| Cost per hospitalization episode – without severe outcome ^27^ | 65-74 | 3,775.21 € | 3,707.99 € | 5,301.32 € |
|  | 75+ | 3,887.45 € | 3,812.08 € | 4,918.05 € |
| Cost per hospitalization episode – with severe outcome ^27^ | 65-74 | 8,978.41 € | 8,669.59 € | 11,292.22 € |
|  | 75+ | 5,192.05 € | 5,335.55 € | 7,855.57 € |

**Abbreviations**: aQIV, adjuvanted quadrivalent vaccine; HD-QIV, high dose quadrivalent vaccine; hosp., hospitalization; LoS, length of stay; P&I, pneumonia and influenza; rVE, relative vaccine efficacy; SD-QIV, standard dose quadrivalent vaccine; VE, absolute vaccine efficacy.

**Supplementary Table 3.** Results from the scenario analysis

|  | **aQIV** | **HD-QIV** | **Incremental** |
| --- | --- | --- | --- |
| **P&I hospitalizations – rVE for aQIV 0%** | | | |
| Total costs | 22,950,842,428 € | 22,930,827,922 € | -20,014,506 € |
| Total QALYs | 88,785,928 | 88,797,914 | 11,986 |
| ICER (€/QALY) |  | HD-QIV dominates aQIV | |
| **P&I hospitalizations – rVE for aQIV 20.0%** | | | |
| Total costs | 22,905,832,050 € | 22,930,827,922 € | 24,995,872 € |
| Total QALYs | 88,786,264 | 88,797,914 | 11,650 |
| ICER (€/QALY) |  |  | 2,146 €/QALY |
| **Respiratory hospitalizations – rVE for aQIV 0%** | | | |
| Total costs | 23,163,264,695 € | 23,145,228,270 € | -18,036,425 € |
| Total QALYs | 88,784,297 | 88,796,273 | 11,975 |
| ICER (€/QALY) | HD-QIV dominates aQIV | | |
| **Respiratory hospitalizations – rVE for aQIV 7.0%** | | | |
| Total costs | 23,139,022,260 € | 23,145,228,270 € | 6,206,010 € |
| Total QALYs | 88,784,480 | 88,796,273 | 11,792 |
| ICER (€/QALY) |  |  | 526 €/QALY |
| **All-cause hospitalizations – rVE for aQIV 0%** | | | |
| Total costs | 30,537,983,331 € | 30,195,350,828 € | -342,632,504 € |
| Total QALYs | 88,744,620 | 88,758,298 | 13,678 |
| ICER (€/QALY) | HD-QIV dominates aQIV | | |
| **All-cause hospitalizations – rVE for aQIV 6.0%** | | | |
| Total costs | 30,229,348,927 € | 30,195,350,828 € | -33,998,099 € |
| Total QALYs | 88,746,335 | 88,758,298 | 11,963 |
| ICER (€/QALY) | HD-QIV dominates aQIV | | |

**Abbreviations**: aQIV, adjuvanted quadrivalent vaccine; HD-QIV, high dose quadrivalent vaccine; ICER, incremental cost-effectiveness ratio; P&I, pneumonia and influenza; QALY, quality-adjusted life year; SD-QIV, standard dose quadrivalent vaccine.
